# Increased peripheral blood neutrophil activation phenotypes and NETosis in critically ill COVID-19 patients

**DOI:** 10.1101/2021.01.14.21249831

**Authors:** Jorge A. Masso-Silva, Alexander Moshensky, Michael T. Y. Lam, Mazen Odish, Arjun Patel, Le Xu, Emily Hansen, Samantha Trescott, Celina Nguyen, Roy Kim, Katherine Perofsky, Samantha Perera, Lauren Ma, Josephine Pham, Mark Rolfsen, Jarod Olay, John Shin, Jennifer M. Dan, Robert Abbott, Sydney Ramirez, Thomas H. Alexander, Grace Y. Lin, Ana Lucia Fuentes, Ira Advani, Deepti Gunge, Victor Pretorius, Atul Malhotra, Xin Sun, Jason Duran, Shane Crotty, Nicole G. Coufal, Angela Meier, Laura E. Crotty Alexander

**Affiliations:** Pulmonary and Critical Care Section, VA San Diego Healthcare System, La Jolla, CA 92161; Division of Pulmonary, Critical Care and Sleep Medicine, Department of Medicine, University of California San Diego (UCSD), La Jolla, CA 92093; The Salk Institute, La Jolla, CA; Rady Children’s Hospital, San Diego, CA; Division of Infectious Disease, Department of Medicine, UCSD, La Jolla, CA 92093; La Jolla Institute of Allergy and Immunology, La Jolla, CA; Division of Head & Neck Surgery, Scripps Clinic, La Jolla, CA; Department of Pathology, UCSD, La Jolla, CA 92093; Division of Cardiovascular and Thoracic Surgery, Department of Surgery, UCSD, La Jolla, CA 92093; Department of Pediatrics, UCSD, La Jolla, CA 92093; Division of Cardiology, Department of Medicine, UCSD, La Jolla, CA 92093; Department of Anesthesiology, Division of Critical Care, UCSD, La Jolla, CA 92093

**Author notes:** authors contributed equally (Jorge A. Masso-Silva, Alexander Moshensky). co-corresponding authors: Angela Meier and Laura Crotty Alexander 3350 La Jolla Village Dr, MC 9111J, San Diego, CA 92161.

## Abstract

**Background:** Increased inflammation is a hallmark of COVID-19, with pulmonary and systemic inflammation identified in multiple cohorts of patients. Definitive cellular and molecular pathways driving severe forms of this disease remain uncertain. Neutrophils, the most numerous leukocytes in blood circulation, can contribute to immunopathology in infections, inflammatory diseases and acute respiratory distress syndrome (ARDS), a primary cause of morbidity and mortality in COVID-19. Changes in multiple neutrophil functions and circulating cytokine levels over time during COVID-19 may help define disease severity and guide care and decision making.

**Methods:** Blood was obtained serially from critically ill COVID-19 patients for 11 days. Neutrophil oxidative burst, neutrophil extracellular trap formation (NETosis), phagocytosis and cytokine levels were assessed *ex vivo*. Lung tissue was obtained immediately post-mortem for immunostaining.

**Results:** Elevations in neutrophil-associated cytokines IL-8 and IL-6, and general inflammatory cytokines IP-10, GM-CSF, IL-1b, IL-10 and TNF, were identified in COVID-19 plasma both at the first measurement and at multiple timepoints across hospitalization (p < 0.0001). Neutrophils had exaggerated oxidative burst (p < 0.0001), NETosis (p < 0.0001) and phagocytosis (p < 0.0001) relative to controls. Increased NETosis correlated with both leukocytosis and neutrophilia. Neutrophils and NETs were identified within airways and alveoli in the lung parenchyma of 40% of SARS-CoV-2 infected lungs. While elevations in IL-8 and ANC correlated to COVID-19 disease severity, plasma IL-8 levels alone correlated with death.

**Conclusions:** Circulating neutrophils in COVID-19 exhibit an activated phenotype with increased oxidative burst, NETosis and phagocytosis. Readily accessible and dynamic, plasma IL-8 and circulating neutrophil function may be potential COVID-19 disease biomarkers.

## Introduction

Since the appearance of SARS-CoV-2 in Wuhan, China in December 2019, to the moment that the World Health Organization (WHO) declared COVID-19 a pandemic on March 11^th^, 2020 and beyond, there has been an imperative to understand better disease pathogenesis to develop effective treatments and vaccines. Acute respiratory distress syndrome (ARDS) and systemic inflammation characterize severe forms of the disease. Similar to SARS, pulmonary failure in SARS-CoV-2 infection is due to both viral and host factors, including exacerbated inflammation [1, 2].

Detrimental host immune responses to infections (immunopathology) are widely recognized and studied in respiratory infections and inflammatory diseases [3-5]. Immunopathology in the context of respiratory viral infections involves increased activity of innate immune cells, including natural killer (NK), NK T-cells (NKT), innate lymphoid cells (ILC), inflammatory monocytes, macrophages and neutrophils [4]. Among these cells, neutrophils are potent early mediators of innate immune responses during infection available to clear pathogens but sometimes at a cost of collateral inflammatory tissue damage [4, 6]. Neutrophil recruitment and their robust activation increase lung tissue damage [6-8]. Recently, neutrophils have been proposed to play a role in acute lung injury (ALI) and ARDS related to COVID-19 [9-11], based on the concept that local neutrophil recruitment and activation leads to hypoxemic respiratory failure [12]. While several groups have highlighted this potential detrimental effect of neutrophil activation in COVID-19 disease pathology [9-11, 13], no in-depth functional assessments of neutrophils isolated from COVID-19 patients exist.

Neutrophils target invading pathogens through various intracellular and extracellular mechanisms, with phagocytosis, production of reactive oxygen species (ROS) and release of neutrophil extracellular traps (NETosis) representing three of the most important mechanisms in their antimicrobial arsenal [14]. Neutrophils produce cytokines and chemokines that contribute to the inflammatory milieu during active infections, and ROS and NETosis have been implicated in mediating the damage to surrounding tissue [6, 15, 16]. In COVID-19, two recent studies associated severe disease with circulating neutrophilia [17, 18], and COVID-19 patient neutrophils showed higher activation marker expression than those of healthy controls [18]. Elevated levels of NET components including myeloperoxidase (MPO), citrullinated histone H3, and DNA are seen in COVID-19 patient sera [18, 19]. While treatment of naïve neutrophils from healthy donors with serum from COVID-19 patients stimulated NETosis [19], and NETs are seen in lung tissue of COVID-19 patients [20], assays have not been on neutrophils isolated from COVID-19 patients to define their functional capacity and association with disease severity.

We sought to define carefully the functional state of neutrophils freshly isolated from the circulation of critically ill COVID-19 patients. By obtaining neutrophils and cytokine profiles from on multiple days over the course of illness, our studies had robust power to detect corresponding changes in neutrophil function and systemic inflammation in this novel disease. Consequently, correlations between circulating leukocytes, neutrophil numbers, plasma IL-8 levels, neutrophil function and disease severity were identified.

## Methods

### Study design and oversight

The research protocol was approved by the UCSD, VASDHS and Rady Children’s Hospital institutional review boards (IRBs) and all participants or designated family member gave written informed consent. Critically ill COVID-19 patients hospitalized at UCSD and Rady Children’s Hospital were recruited for these studies. After informed consent, blood was drawn on days 1, 3, 5, 7, 9, 1, most commonly between 8-10 am, and immediately prior to discharge. On days where 11 ml of blood or greater was obtained, neutrophils were isolated for functional studies. Healthy controls were recruited from VASDHS and UCSD staff members. All healthy control subjects underwent informed consent, passed COVID-19 screening protocols on each blood draw day, and donated blood once a week for four weeks. Rapid autopsy for donation of tissue for research was conducted under a waiver from the UCSD IRB.

## Subjects

Data from all 16 hospitalized COVID-19 patients with neutrophil data obtained on multiple days were included. Data from an additional 4 hospitalized COVID-19 patients were also used for the cytokine studies. APACHE (acute physiology and chronic health evaluation) II scores were calculated using clinical data within 24 h of blood draws for neutrophil functional assays derived by chart review. Other scores were also considered – SOFA (Sequential Organ Failure Assessment) was excluded due to a lack of variable diversity and small range of values. APACHE III and IV are more complex scores with additional variables, but yielded no additional sensitivity to detecting changes in disease severity in this patient population. APACHE II is known to have consistent calibration compared to III and IV, and more subjects globally are assessed with the APACHE II. COVID-19 patients had APACHE II scores ranging from 7 to 27 on ICU admission. Healthy controls were attributed an APACHE score of 0. White blood cell count (WBC), absolute neutrophil count (ANC) and neutrophil:lymphocyte ratios were captured from the clinical data of SARs-CoV-2 positive patients throughout their hospital and were closely matched to the timing of blood draws for neutrophil assays. Lung tissue was obtained within 2 h postmortem from a 67 year-old male COVID-19 patient, intubated for 17 days prior to death in July 2020.

### Cytokine quantification

The Human Anti-Virus Response Panel (13-plex; BioLegend Inc) was used to assess human plasma cytokines (IL-1β, IL-6, IL-8, IL-10, TNFα, GM-CSF and IP-10) from plasma samples obtained across multiple timepoints and stored at ^-^80°C until ready for use. On the day of the assay, plasma was thawed at room temperature, centrifuged at 1,000 g for 5 min, and run at a 2-fold dilution in Assay Buffer using the recommended 96-well filter plate method precisely per the manufacturer’s protocol. Samples were acquired on a Canto II flow cytometer (BD) using a high throughput sampler and run in duplicate unless plasma volume was inadequate with corresponding standards on all plates. Cell signaling technology (CST) was run prior to all flow cytometry runs to ensure low detector CVs and set laser delay. LEGENDplex Data Analysis Software (BioLegend) was used for analysis.

### Neutrophil cell isolation and functional assays

*Neutrophil isolation*: On multiple days across the hospital stay for COVID-19 subjects, and once a week for control subjects, blood was drawn into lithium heparin tubes (BD Vacutainer) with an average gap of 2.5 h to transfer time prior to neutrophil isolation —this gap was mimicked for neutrophil isolation from healthy controls. Neutrophils were isolated using Polymorphprep^TM^ (PROGEN) per manufacturer’s instructions as previously described [21]. Briefly, whole blood was layered on 20 ml of Polymorphprep^TM^ in a 50 ml conical tube and centrifuged at 500 × *g* for 30 min at 20°C, sans brake. The granulocyte layer was collected and washed with HBSS with no calcium, no magnesium (HBSS^-/-^) and centrifuged at 400 × *g* for 10 min at 20°C. The cell pellet was resuspended in 1 ml HBSS^-/-^, 10 uL was cytospun and Giemsa Wright stained, and cells were counted using light microscopy and a hemocytometer. Average neutrophil purity across samples was 90%. Cells were resuspended at 2×10^6^ cells/ml for functional assays.

### NET staining within lung tissue

Lung tissue obtained by rapid bedside autopsy was fixed with 4% paraformaldehyde for 48 h then processed for PBS washing and paraffin embedding. After deparaffinization and rehydration, tissue slides were subjected to antigen retrieval in high PH buffer (Tris-EDTA with 0.05% Tween20, Ph 9.0) at 100°C for 12 min. After blocking with goat or donkey serum, primary antibodies were incubated overnight at 4°C. Incubation of secondary antibodies for 1 h at room temperature was followed by treatment with TrueBlack (Biotium) to reduce autofluorescence. Images were acquired by Zeiss fluorescence microscope (ZEISS Axio Imager 2). Primary antibodies used for immunofluorescence were: CD66b (1:100, 392902, Biolegend), citrullinated H3 (H3-cit) (1:100, ab5103, Abcam), myeloperoxidase (MPO) (1:100, AF3667, R&D), and histone-2B (H2B) (1:100, 606302, Biolegend). 4′,6-diamidino-2-phenylindole (DAPI) was used for nucleic acid staining. Haemotoxylin and Eosin (H&E) staining was conducted by using the standard protocol.

### Statistical analysis

Statistical analyses were performed using GraphPad Prism Version 8.4.3 (San Diego). Patient demographics were compared using the t-test or Fisher’s exact test, as appropriate. Plasma cytokine levels and NETosis in controls and COVID-19 subjects were compared with the Mann-Whitney test. ROS production and phagocytosis assays were analyzed using mixed-effects models with Geisser-Greenhouse correction and Sidak’s multiple comparisons test for each timepoint. Correlation of NETosis with WBC, ANC and APACHE score were done with linear regression. Analyses of APACHE score correlation with ANC and IL-8 were performed with linear regression. Comparison of IL-8 levels between controls and patients with APACHE score groups was performed with ANOVA with Sidak’s correction for multiple comparisons. Two-sided tests and α level of 0.05 were used to determine significance.

## Results

### Characteristics of the COVID-19 patients and healthy controls

Mean age was 57.5 years for COVID-19 patients and 40 years for controls (p = 0.008; Table 1). The COVID-19 cohort included equal numbers of female and male subjects. All COVID-19 patients had pre-existing conditions, with hypertension being the most prevalent (50%), followed by diabetes mellitus (25%), COPD (25%), congestive heart failure (12.5%), ischemic stroke (6%)and asthma (6%).

**Table 1.** Demographics by Group Category.

|  | <u>Controls</u><br>n=13 (%) | <u>COVID-19</u><br><u>Patients</u><br>n=16 (%) | <u>Overall</u><br>n=29 (%) | p value |
| --- | --- | --- | --- | --- |
| **Gender** |  |  |  |  |
| Female | 5 (38.46%) | 7 (43.75%) | 12 (41.38%) | 0.774 |
| Male | 8 (61.54%) | 9 (56.25%) | 17 (58.62%) |  |
| **Race** |  |  |  |  |
| African American | 0 (0.00%) | 0 (0.00%) | 0 (0.00%) | 0.031 |
| Caucasian | 9 (69.23%) | 16 (100.00%) | 25 (86.21%) |  |
| East Asian | 4 (30.77%) | 0 (0.00%) | 4 (13.79%) |  |
| Other | 0 (0.00%) | 0 (0.00%) | 0 (0.00%) |  |
| **Ethnicity** |  |  |  |  |
| Hispanic | 3 (23.08%) | 11 (68.75%) | 14 (48.28%) | 0.025 |
| Non-Hispanic | 10 (76.92%) | 5 (31.25%) | 15 (51.72%) |  |
| **Age** |  |  |  |  |
| Mean (range) | 40 (25 - 53) | 57.5 (17 - 88) | 50.4 (17 - 88) | 0.008 |

Demographic data was taken from healthy controls as well as SARS-CoV-2 positive patients (COVID-19). Categorical variables were compared with chi-squared test. Continuous variables were compared with an unpaired t-test.

Fifteen out of 16 COVID-19 patients were admitted to the ICU (94%) with average APACHE II score 16.5 (range of 7-27). Patients averaged 6 days from first symptom to ICU admission and remained in the ICU for an average of 14 days (range 4-32). The mortality rate for this cohort was 19% (n = 3). All ICU patients required vasopressors and 88% required invasive mechanical ventilation. Ventilated patients had an average Murray score of 2.6 and an average P/F ratio of 128 prior to intubation and 206 at extubation. The average PEEP requirement at intubation was 11 and peak PEEP was 13. Only three patients received steroids. A wide variety of antimicrobials and antivirals were given, with vancomycin, piperacillin-tazobactam, azithromycin and ceftriaxone the most common. Micafungin and acyclovir were used once, and remdesivir was used in 5 patients as part of clinical trials (Supplementary Table 1).

### Increased neutrophil and general inflammatory cytokines in COVID-19

Plasma cytokine profiles and complete blood counts of our COVID-19 patient cohort demonstrated similarities to published cohorts [22-25], with elevations in IL-8, IL-6, IP-10, and of the neutrophil:lymphocyte ratio (mean 9.3). Profiling of specific cytokines relevant to neutrophil activity showed broad elevations across IP-10, IL-6, IL-8, GM-CSF, IL-1β, IL-10 and TNFα in the circulation of critically ill COVID-19 patients both early in their hospitalization (Fig. 1A) and were persistently elevated across their hospitalization, assessed at multiple time points (Fig. 1B).

**Figure 1.**
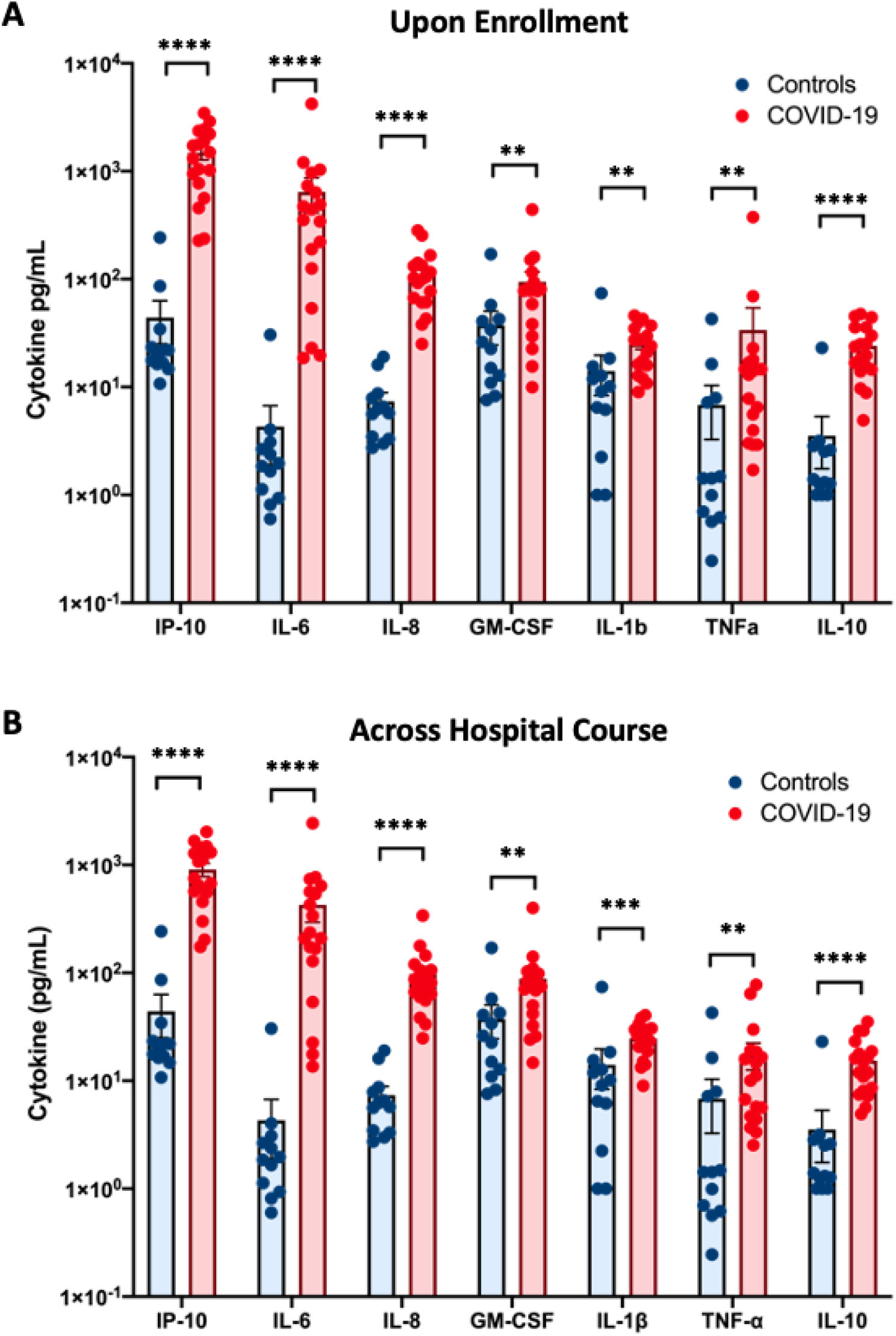
Pro-inflammatory cytokines are increased in the plasma of critically ill COVID-19 patients. Consistent with prior reports, cytokines in the circulation of COVID-19 patients were elevated at the first timepoint assessed (**A**) and remained elevated persistently across their hospital courses (**B**). At the time of enrollment in the study, most typically at the time of ICU admission, all cytokines were significantly elevated relative to controls at this first timepoint (**A**). IP-10, IL-6, IL-8, GM-CSF, IL-1β, IL-10 and TNFα were elevated in the plasma of COVID-19 patients (n= 16) when averaged across all timepoints tested, relative to healthy controls (n = 13; **B**). Error bars represent standard error of the mean. **p < 0.01, ***p < 0.001, ****p < 0.0001.

### Circulating neutrophils from COVID-19 patients have higher rates of NETosis than healthy individuals

Circulating neutrophils receive inflammatory signals prior to entering tissues that impact their potential detrimental activity once in the tissue. There is mounting evidence of the role of NETs in systemic and pulmonary inflammation in COVID-19 [20, 26]. We investigated NET production by circulating neutrophils *ex vivo* by relative fluorescence of released dsDNA that may parallel exacerbated inflammation in the lungs. Neutrophils isolated from COVID-19 patients produced more NETs in steady state (without stimulation) than neutrophils from healthy individuals, and such differences were consistent even upon stimulation with different concentrations of PMA (Fig. 2A). We observed the same trend when using fluorescence microscopy staining for myeloperoxidase, a major protein component of NETs (Fig. 2B). These data show that circulating neutrophils from COVID-19 patients are more prone to produce NETs than neutrophils from healthy individuals, and thus may be primed to cause significant NET-mediated tissue injury once they enter into lung tissue.

**Figure 2.**
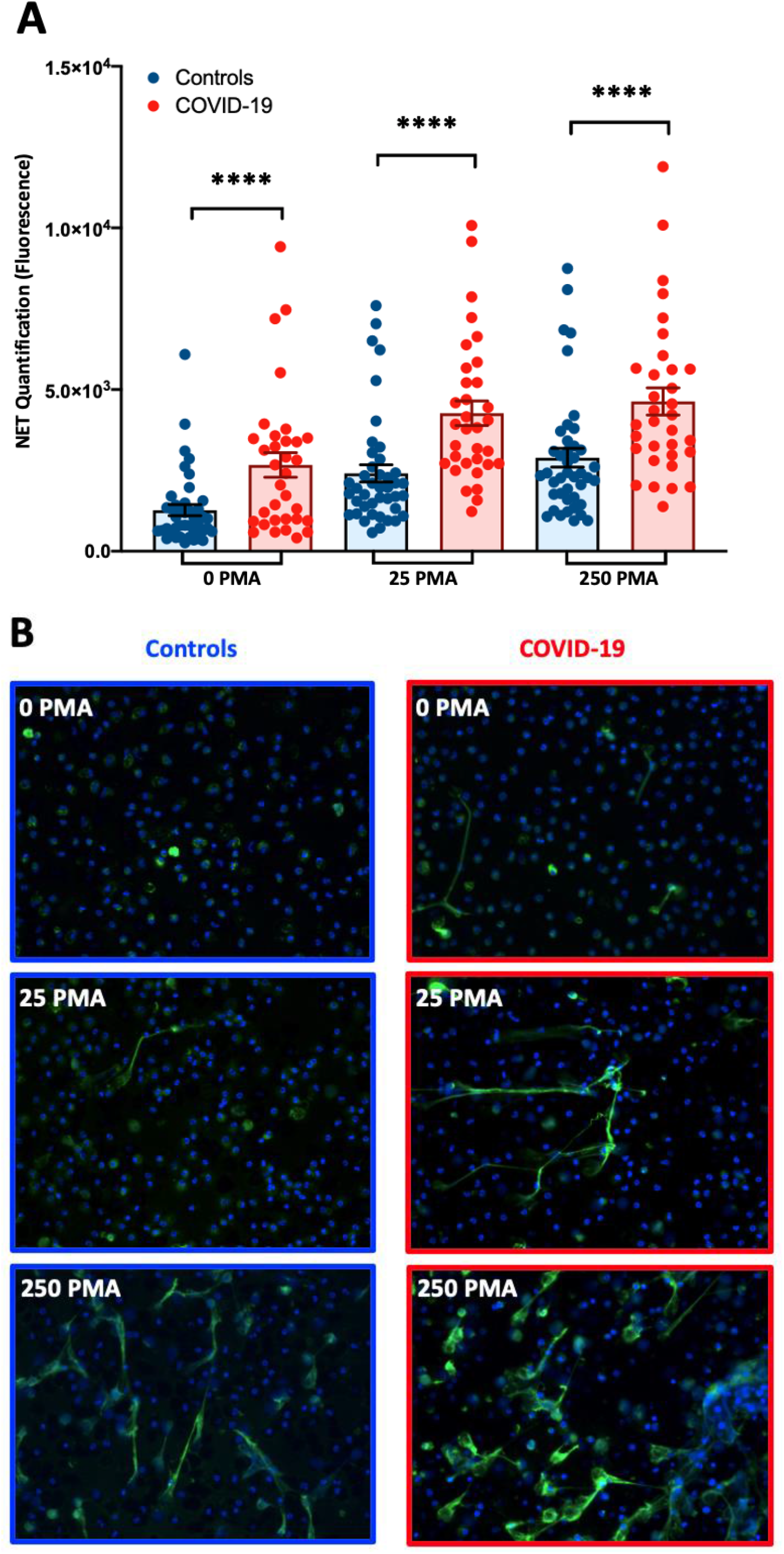
Increased NETosis by circulating neutrophils from COVID-19 patients. Functional NETosis assays on neutrophils isolated from the blood of critically ill COVID-19 patients on multiple days across their hospitalization (n = 16 subjects; total timepoints represented = 33) and healthy controls over time (n = 13; total timepoints represented = 42) demonstrate that neutrophils from COVID-19 patients have higher NETosis at steady state and also upon stimulation with PMA (both 25 and 250ug), assessed by A) quantification of dsDNA by fluorescence, and B) fluorescence microscopy of neutrophils stained for Myeloperoxidase (green) and DNA (blue). Error bars represent standard error of the mean. ****p < 0.0001.

### Neutrophils and NETs present in terminal bronchioles and alveoli of COVID-19 lung tissue

Beside their antimicrobial function, release of NETs has been associated with collateral host tissue damage, including the lungs [10, 15]. Deterioration of lung function due to cellular damage and inflammation with subsequent hypoxemia is a hallmark feature of severe COVID-19, although the underlying mechanisms remain unknown [27]. To investigate the presence of neutrophils and NETs in COVID-19 lung tissue, we performed rapid autopsies on five deceased patients, plus additional non-COVID controls, which demonstrated a multitude of neutrophils across airways and alveoli by H&E staining of lung tissue in two of the five patients (Figure 3A). Specifically, alveolar lung parenchyma showed extensive organizing pneumonia with areas of interstitial fibrosis, consistent with prior diffuse alveolar damage, with the majority of neutrophils within the terminal/respiratory bronchioles with focal areas with neutrophils within the alveolar lung parenchyma (Figure 3A). Bronchiolar metaplasia was also seen. Positive staining for citrullinated Histone H3, CD66b, and DAPI confirmed the presence of neutrophils and NETs within bronchioles, alveoli and lung parenchyma (Figure 3B); NETs were furthermore positive for MPO (Supplemental Figure 1). In the two SARS-CoV-2 infected lungs found to have NETs, the majority of NETs were found within terminal respiratory bronchioles, with focal areas within the alveolar lung parenchyma.

**Figure 3.**
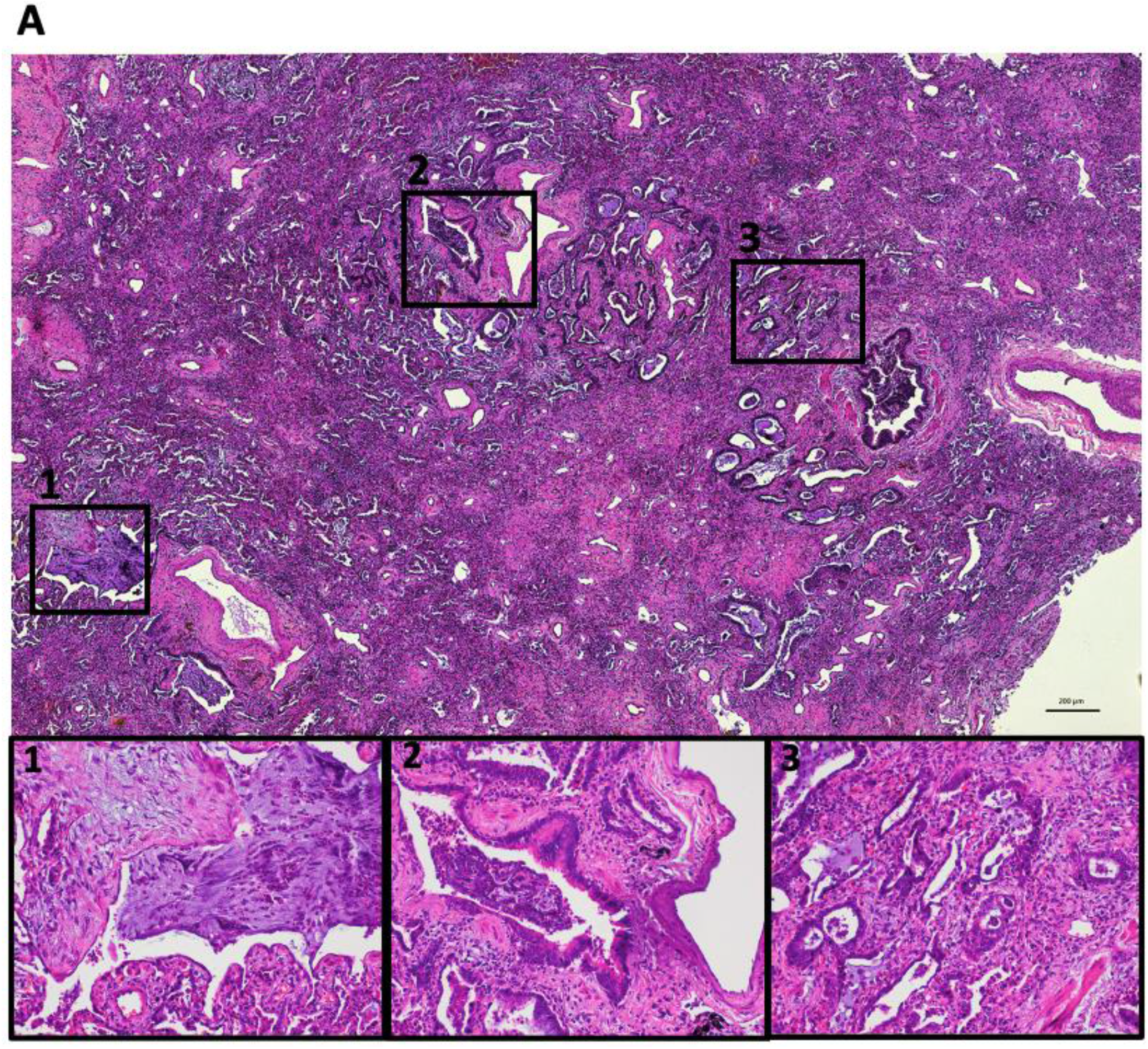
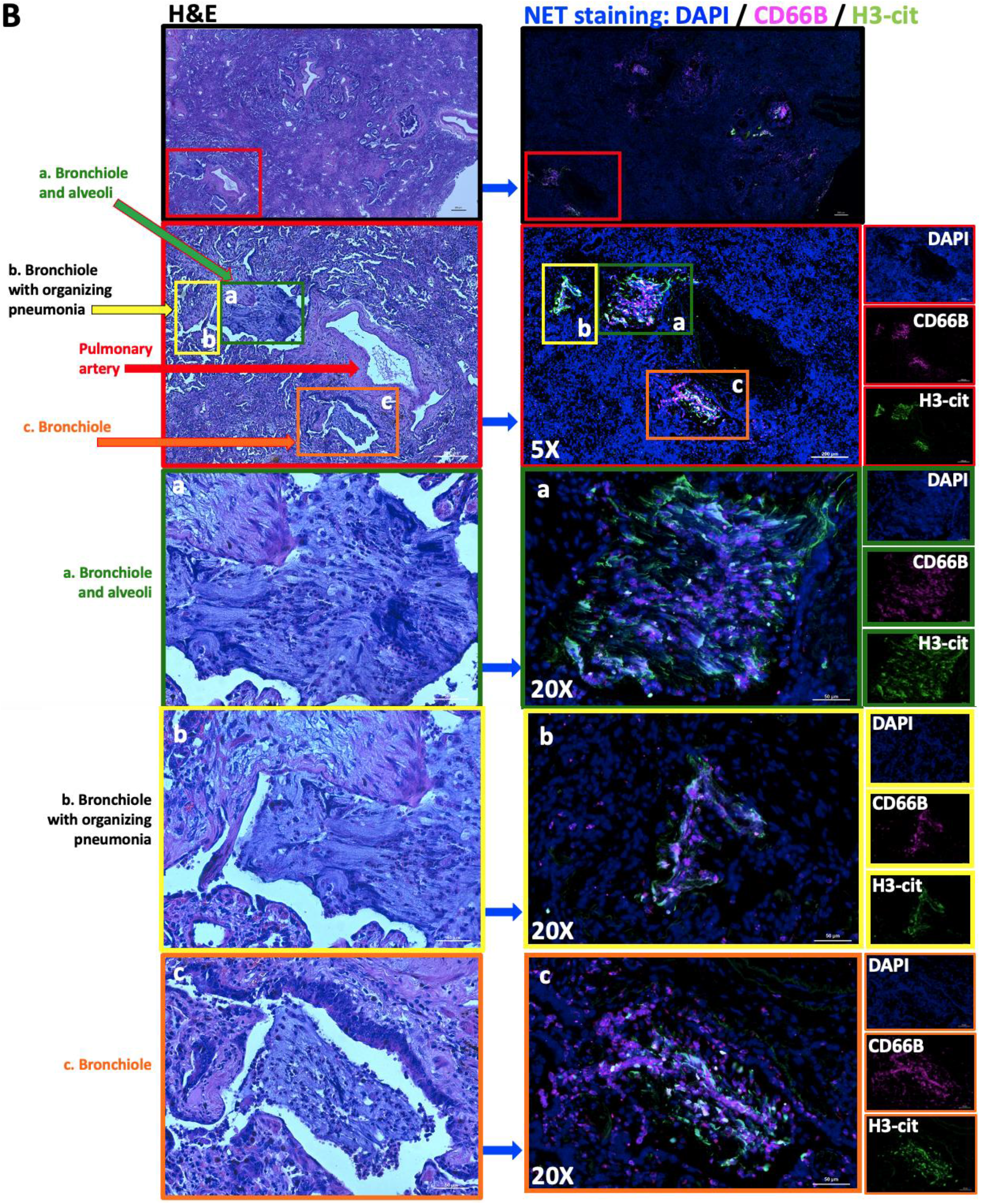
Neutrophils and NETs throughout airways and parenchyma of COVID-19 lung tissue. **A**) H&E staining demonstrates diffuse inflammation and fibrosis with the presence of neutrophils throughout alveoli and bronchioles. The majority of neutrophils were found within terminal bronchioles, with focal areas of neutrophils within the alveolar lung parenchyma. 1. Terminal bronchiole and alveoli with neutrophils throughout. 2. Neutrophils within fibrotic lung parenchyma with bronchiolar metaplasia. 3. Neutrophils present throughout areas of organizing pneumonia. **B**) Immunofluorescence staining of nuclei with DAPI (blue), CD66b (magenta) and H3-cit (green) demonstrate the presence of neutrophils and NETs within the lung parenchyma. Bronchioles, alveoli and pulmonary arteries are indicated with colored arrows and squares. Three areas of NETs are highlighted and shown at 20x magnification, with H&E images for reference.

### Circulating neutrophils from COVID-19 patients have increased production of reactive oxygen species and increased phagocytosis

ROS production is an antimicrobial mechanism mechanistically linked to NETosis and known to cause tissue damage [28]. Mirroring the NETosis phenotype, ROS production was increased in COVID-19 patients as compared to healthy controls at steady state over time (no stimulation; Fig. 4A), and functional capacity increased upon stimulation with PMA at 2.5 nM (Fig. 4B), 25 nM (Fig. 4C) and 250 nM (Fig. 4D). Circulating neutrophils from COVID-19 patients also demonstrated increased phagocytosis of *S. aureus* bioparticles (Fig. 4E), suggesting that multiple key innate antimicrobial and inflammatory functions of circulating neutrophils patients are significantly increased in COVID-19 patients.

**Figure 4.**
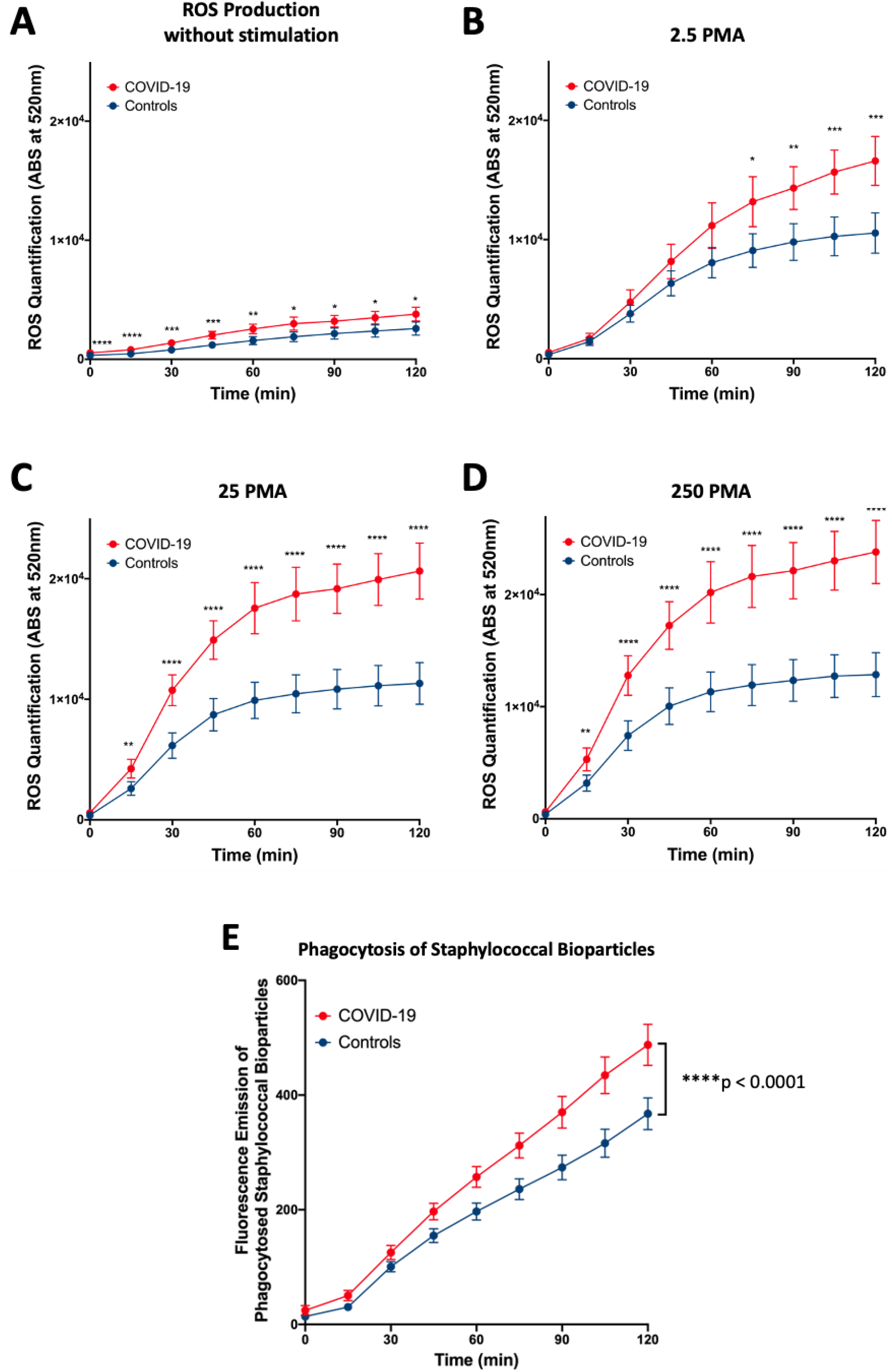
Circulating neutrophils from COVID-19 patients produce more reactive oxygen species and have increased phagocytosis than those from healthy subjects. Reactive oxygen species (ROS) production and phagocytosis by neutrophils isolated from the blood of COVID-19 patients and healthy controls were assessed by relative fluorescence. ROS production was significantly higher at A) steady state, B) 2.5 nM PMA, C) 25 nM PMA and D) 250 nM PMA. E) Phagocytosis was significantly higher in COVID-19 neutrophils relative to neutrophils from healthy controls. Data was analyzed with mixed-effects models with Geisser-Greenhouse correction, giving p < 0.0001 for all analyses. Error bars represent 95% CI. ^*^p < 0.05, ^**^p < 0.01, ^***^p < 0.001, ^****^p < 0.0001.

### Increased NETosis is associated with leukocytosis and neutrophilia, and leukocytosis, neutrophilia and plasma IL-8 correlate with COVID-19 disease severity

To assess absolute neutrophil count (ANC) and leukocytosis as potential biomarkers for disease severity in COVID-19, we assessed the relationship between WBC and ANC with our *ex vivo* data for NETosis at steady state (without stimulation) while controlling for neutrophil numbers in our experiments. We found a positive correlation of WBC to NETosis (Fig. 5A). Patients with leukocytosis (WBC greater than 11×10^3^ cells/mm^3^) produced more NETs per neutrophil than patients without leukocytosis (r^2^ = 0.48, p <0.0001; Fig. 5B). Similarly, we found a positive correlation of ANC to NETosis (r^2^ = 0.53, p <0.0001; Fig. 5C) and patients with neutrophilia (ANC greater than 8×10^3^ cells/mm^3^) produced more NETs per neutrophil than patients without neutrophilia (Fig. 5D). Higher levels of neutrophils correlated with disease severity, as measured by APACHE II (r^2^ = 0.24, p = 0.005; Fig. 5E), while lymphocyte counts fell as disease severity increased (r^2^ = 0.19, p < 0.001; Fig 5F). These data demonstrate that COVID-19 patients with leukocytosis, neutrophilia and lymphopenia are highly likely to have increased inflammatory and antimicrobial functions, including NETosis.

**Figure 5.**
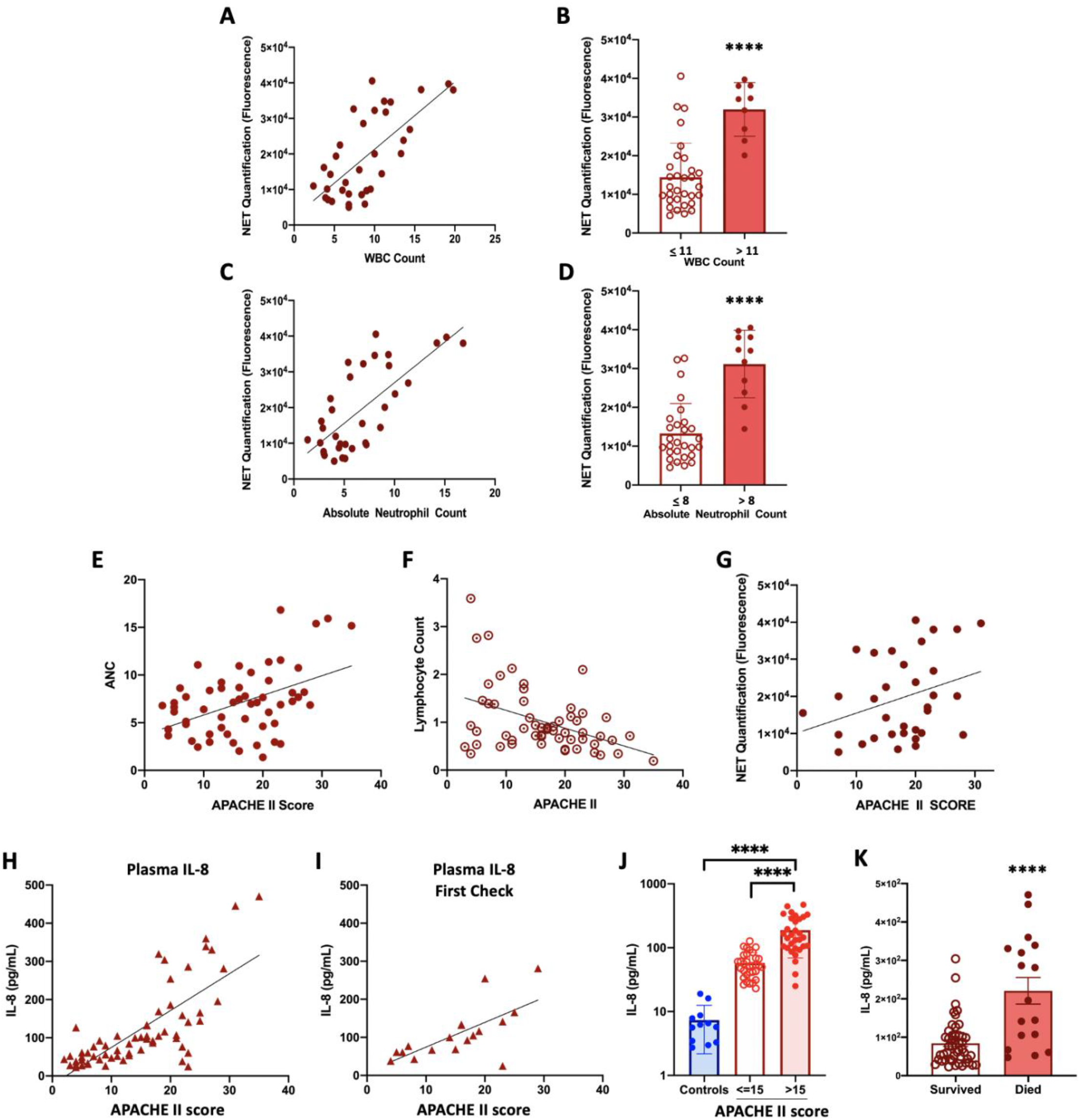
Increased NETosis is positively correlated with leukocytosis and neutrophilia, and neutrophilia and IL-8 are positively correlated with COVID-19 severity. Levels of NETosis were correlated to white blood cell counts (WBC) and absolute neutrophil counts (ANC) from COVID-19 patients. A) Scatter plot demonstrated positive correlation of WBC and NETosis (r^2^ = 0.48, p<0.0001). B) Patients with leukocytosis produced significantly more NETs than patients with WBC levels under 11×10^3^/mm^3^. C) Scatter plot showed positive correlation of ANC and NETosis (r^2^ = 0.53, p <0.0001). D) Patients with neutrophilia produced significantly more NETs than patients with ANC levels under 8×10^3^/mm^3^. E) Scatter plot showing that severity of COVID-19 positively correlated with neutrophilia (r^2^ = 0.24, p = 0.005). F) Decreasing lymphocyte counts were found as COVID-19 severity rose. G) NETosis was not found to correlate with severity of disease (r^2^ = 0.11, p = 0.069). H) Severity of COVID-19 correlated with IL-8 levels quantified on multiple days across hospitalization (r^2^ = 0.403, p = 0.006). I) IL-8 levels at the first time point, on entry to the study, also correlated with disease severity (r^2^ = 0.449, p = 0.034). J) Plasma IL-8 levels were highest in COVID-19 patients with the greatest disease severity (APACHE II >15). K) Patients who succumbed to their disease had greater IL-8 plasma levels than those who survived. Error bars represent 95% CI. ^****^p < 0.0001.

We also explored a potential direct relationship between NETosis and disease severity, using the APACHE II score as an indicator for illness severity, but found no such correlation (r^2^ = 0.10, p = 0.07; Fig. 5G). To be thorough, we also assessed the relationship of NETosis with SOFA and COVID-GRAM scores, and found no associations. However, plasma levels of IL-8 significantly correlated with higher APACHE II scores, both across hospitalization (Fig. 5H) and on the first check (Fig. 5I). All other elevated cytokines were also assessed for a disease severity correlation, but none was found. The neutrophil-associated cytokine was highest in the plasma of the most critically ill patients as compared to healthy controls and those less severely ill (Fig. 5J), and IL-8 was significantly higher in those who succumbed to the disease (Fig. 5K).

## Discussion

The COVID-19 pandemic has struck the world affecting many aspects of life, and we are undoubtedly in a race against time to find effective treatments and vaccines against SARS-CoV-2. Our work focused on examining the role of neutrophil activity in COVID-19 patients, due to the many roles of these cells in lung and systemic tissue damage in the context of infectious and non-infectious challenges [29]. The serial measurements (with pairing of clinical state with neutrophil function and cytokine levels on the same day) allowed more in-depth assessment of the immune state in COVID-19, and allowed for assessment of the roles of many clinical factors in neutrophil function. We particularly looked at medications used and treatment modalities applied and did not find any that were significantly associated with neutrophil functional changes. We confirmed the presence of neutrophils and NETs within alveoli and airways within lung tissue from two deceased COVID-19 patients, which complements recent data whole tissue transcriptomic analysis of lung tissue and bronchoalveolar lavage fluid (BALF) that found upregulation of genes associated with NETosis [17], and recent data on the presence of NETs within lung tissue of COVID-19 patients [20]. However, the absence of NETs within lung parenchyma in three deceased COVID-19 patients suggests that there is heterogeneity in the patterns of immunopathology in this disease. This fits well with the current concept of different subtypes of ARDS [30], and confirms that not all COVID-19 ARDS is the same [31].

By assessing *ex vivo* key functional neutrophil activities, we show that circulating neutrophils in COVID-19 have multiple functional changes, including increased oxidative burst, NETosis and phagocytosis. These findings are in alignment with a recent study showing that neutrophils from COVID-19 patients have higher expression of CD66b and decreased CD62L [18], consistent with an enhanced neutrophil activation phenotype. Healthy neutrophils exposed to serum from COVID-19 patients have increased NETosis as compared to neutrophils treated with serum from healthy individuals [19], other studies found NET components in serum and/or plasma of COVID-19 patients, including cell free-DNA [18, 19], MPO-DNA [18, 19], NE-DNA [18] and citrullinated histone H3 [18, 19], and Veras et al. demonstrated increased NETosis in COVID-19 neutrophils [26].

The link between neutrophils and COVID-19 pathogenesis continues to grow stronger, with several reports addressing the potential relevance of these cells and their activities in disease progression [10, 11, 13, 16, 32]. COVID-19 patients have increased frequencies or total numbers of circulating neutrophils compared to healthy individuals [17, 18], and we confirm the leukocytosis and neutrophilia in severe COVID-19, while further showing that both indices are associated with an ability of circulating neutrophils to produce higher amounts of NETs *ex vivo* in steady state and following stimulation. We had hypothesized that neutrophil function might be a potential biomarker in COVID-19, but found a stronger correlation with the neutrophil cytokine IL-8.

While “cytokine storm” has been characteristic of severe forms of COVID-19 [33], plasma IL-6, IL-8 or TNF-α levels at the time of hospitalization have not proven to be strong and independent predictors of patient survival [34]. In our cohort, we confirmed elevated levels of IL-8, IL-6, and IP-10 in the plasma of COVID-19 patients as compared to healthy controls, with IL-8 alone positively correlated with increased severity of disease (assessed by APACHE II scores). These data support the idea of IL-8, the most powerful neutrophil chemokine which also activates oxidative burst in neutrophils, as a potential dynamic biomarker in this novel disease. Moreover, other cytokines were elevated in COVID-19 patients and merit further investigation in the future such as IP-10, GM-CSF, IL-1β and IL-10. While all cytokines were elevated in COVID-19 subjects across their hospitalizations, longitudinally, each cytokine was also significantly elevated at the first check. These data suggest that cytokine profiling upon admission may help detect patients at higher risk for severe disease.

Our study has limitations. Our control subjects were younger, had higher numbers of Asian ancestry and fewer of Hispanic heritage than our COVID-19 patients. This situation is primarily due to the limitation to conduct human subjects research during the COVID-19 pandemic, such that we were limited to recruiting from employees at the VA San Diego Healthcare System. These control subjects were primarily to define the range of normal for neutrophil functional assays, run on different days, as there is significant day to day variability in these assays. Studying neutrophils from these control subjects on multiple days proved that it is feasible to define the activation state of neutrophils at a single timepoint as well as over time. It is unknown whether hypertension, cardiac disease, and ancestry impact neutrophil function. Diabetes and increasing age are associated with diminished neutrophil function [35-37]; thus, we do not believe that the increased neutrophil function in the COVID cohort is due to higher levels of these comorbidities. Another limitation is the size of our cohort, but by obtaining neutrophils, plasma and clinical data at multiple points over time, this approach increased the power of our study to detect clinically significant changes.

The role of neutrophils in COVID-19 immunopathology is gaining interest rapidly. In aggregate, ours is the first study to report broad functional assessments of circulating neutrophils isolated from critically ill COVID-19 patients, and to put these findings in context of plasma cytokine levels, disease severity, and neutrophil counts. We confirm the presence of NETs within the lungs of COVID-19 patients, and support a prior report of NETs within the terminal bronchioles and alveoli. Our data show overall enhanced NET production, ROS production and phagocytic activity, and correlations of WBC, ANC and IL-8 levels with severity of disease, highlighting the potential detrimental contribution of hyperactivates neutrophils during COVID-19. These data support the ongoing efforts to therapeutically target neutrophil-related mechanisms and propose neutrophil-centric biomarkers to utilize in the continuing battle against this devastating disease.

## Data Availability

The data that support the findings of this study are available on request from the corresponding authors (LCA and AM).

## Acknowledgements

This work would not have been possible without numerous staff members at the University of California San Diego, Rady Childrens Hospital (RCH), the VA San Diego Healthcare System, and the Salk Institute. A special thank you to the IRB members at all institutions, to Victor Nizet, MD, for his valuable insight, and to Jennifer Foley, RN, the RCH PICU Research nurse who helped with consents and documentations.

## Sources of Funding

This work was supported by the Department of Veterans Affairs (salary support and VA Merit Award, PI Crotty Alexander) and NIH NHLBI (PI Crotty Alexander).

## Competing Interests Statement

The authors report no conflicts of interest.

## Author Contributions

The study was designed by LCA, AM, JAMS, AM, NC, ML and JD. The experiments were performed by JAMS, AM, JO, JD, LCA, VP and LX. The data were analyzed by JMS, AM, LCA, SP, LM, JP, AP, JAMS, ALF, JO, TA, XS, ALF and GL. The manuscript was written by LCA, JAMS, SC, TA, AM and AP.

## Notes

### Competing Interest Statement

The authors have declared no competing interest.

### Author Declarations

The research protocol was approved by the UCSD, VASDHS and Rady Childrens Hospital IRBs.

