## Supplemental Data for "Increased peripheral blood neutrophil activation phenotypes and NETosis in critically ill COVID-19 patients"

### Supplemental Table 1. Patient Clinical Course

| | COVID-19 Patients n=16 (%/Range) |
|---|---|
| **Intensive Care Unit (ICU)** | |
| Admitted to ICU | 15 (93.75%) |
| Average APACHE II[a] Score at ICU Admission (range 0-71) | 16.5 (7-27) |
| Average Days from First Symptom to ICU Admission | 6.2 (0-16) |
| Average Days in ICU | 14 (4-32) |
| Vasopressor Requirement | 15 (93.75%) |
| **Respiratory Support** | |
| Maximum Respiratory Support at Admission | |
| Invasive Mechanical Ventilation (IMV) | 11 (68.75%) |
| Non-Invasive Mechanical Ventilation (NIMV) | 1 (6.25%) |
| Non-Rebreather Mask | 2 (12.50%) |
| High Flow Nasal Canula | 2 (12.50%) |
| IMV During Admission | 14 (87.5%) |
| Average Murray[b] Score Prior to Intubation (range 0-4) | 2.6 (1.8-3.0) |
| Average P/F Ratio at Intubation | 127.66 (36-217) |
| Average P/F Ratio at Extubation | 206.16 (77-270) |
| Average PEEP Requirement at Intubation | 11.23 (8-16) |
| Peak PEEP during Admission | 13.31 (8-18) |
| **Steroids** | |
| Dexamethasone | 2 (12.50%) |
| Prednisone[d] | 1 (6.25%) |
| **Antimicrobials/Antivirals** | |
| Azithromycin | 8 (50.00%) |
| Cefepime | 3 (18.75) |
| Ceftriaxone | 7 (43.75%) |
| Meropenem | 4 (25.00%) |
| Micafungin | 1 (6.25%) |
| Vancomycin | 10 (62.50%) |
| Piperacillin-Tazobactam | 9 (56.25%) |
| Aztreonam | 1 (6.25%) |
| Metronidazole | 1 (6.25%) |
| Acyclovir | 1 (6.25%) |
| Remdesivir[e] | 5 (31.25%) |
| **Trials** | |
| Remdesivir vs. Placebo | 1 (6.25%) |

| | |
|---|---|
| Tocilizumab vs. Placebo | 5 (31.25%) |
| Baricitinib vs. Placebo | 4 (25.00%) |
| Received Remdesivir[f] | 5 (31.25%) |
| **Patient Outcome** | |
| Recovered | 13 (81.25%) |
| Death | 3 (18.75%) |

Clinical course of SARs-CoV 2 Patients. Information of the sixteen patients gathered using chart review. APACHE II[a] scored individually using mdcalc.com, APACHE has a range from 0-71 indicating mortality risk within the ICU, 71 being the highest score. Murray[b] score indicates level of acute lung injury prior to intubation, the score was gathered from the EMR with a range from 0-4, 4 being the highest score. Patient given prednisone was due to Lupus Nephritis[d]. Remdesivir[e] within antimicrobials includes all patients which were confirmed to have been given the drug rather than a part of the Remdesivir/placebo trial. All but one patient given Remdesivir were also on the Barcitinib/placebo trial[f].

**Supplemental Figure 1.** Immunofluorescence staining of nuclei with DAPI (blue) and MPO (magenta) demonstrate the presence of NETs within COVID-19 lung parenchyma at both 5x (red boxes) and 20x magnification (green, yellow and orange boxes). (a) MPO staining within a bronchiole and alveoli (green). (b) MPO staining within a bronchiole in an area of organizing pneumonia (yellow). (c) MPO staining within a bronchiole (orange).

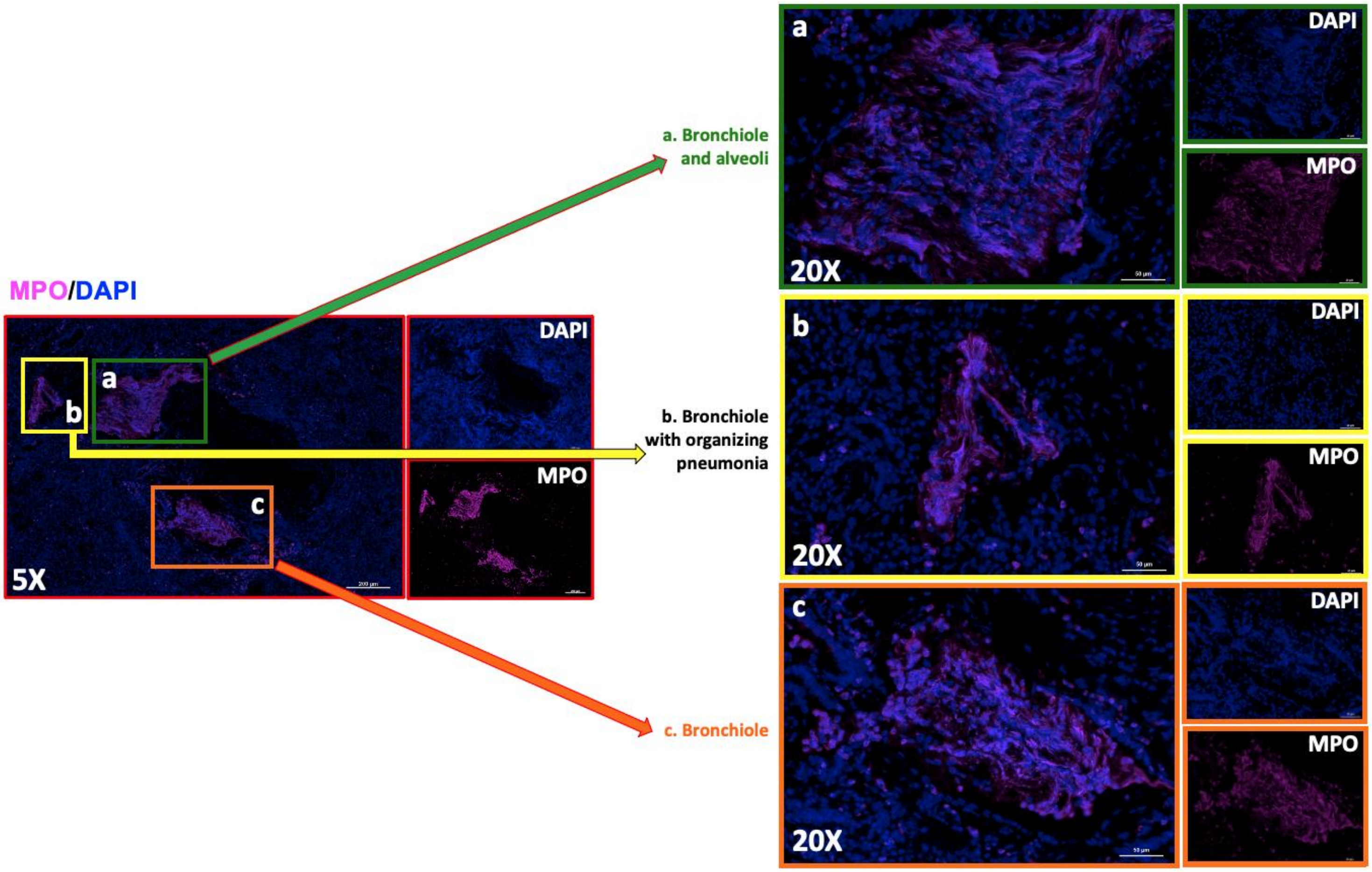

**Supplemental Methods**

*NETosis*: Neutrophils ($2x10^5$ in 100 µl) were plated in a 96-well plate and incubated at 37°C with 5% $CO_2$ with and without phorbol 12-myristate 13-acetate (PMA) at final concentrations of 2.5 nM, 25 nM, and 250 nM (all conditions were run in triplicate). After 2 h, 500 mU/ml of micrococcal nuclease was added to each well and incubated for 10 min at 37°C with 5% $CO_2$. Next, 2.5 µl of 0.5M EDTA was added and cells were centrifuged at 200 x *g* for 8 min. Supernatant (100 µl) was mixed with 100 µl of Quant-iT™ PicoGreen™ (dsDNA Assay Kit, Invitrogen), incubated for 5 min at room temperature in the dark, and fluorescence measured using the Infinite M200 (TECAN) plate reader with 480 nm excitation and 520 nm emission.

*NET imaging*: Neutrophils at $2x10^5$ in 100 µl were plated in wells of an 8-chamber glass slide (Nunc™ Lab-Tek™ II Chamber Slide™ System) and incubated at 37°C with 5% $CO_2$ with 0, 2.5, 25 and 250 nM PMA (all conditions were run in duplicate). After 2 hours, 16% PFA was added to a final concentration of 4% and slides were incubated at 4°C overnight. Supernatants were replaced with 1x PBS and slides were stored at 4°C until staining and visualization. Blocking was conducted for 45 min with 2% bovine serum albumin (BSA) and 2% goat serum in PBS. NETs and neutrophils were immunostained for 1 h with rabbit anti-human myeloperoxidase primary antibody (1:300 in 2% PBS-BSA; Dako North America, Inc.), 45 min with Alexa Fluor 488 goat anti-rabbit IgG secondary antibody (1:500 in 2% PBS-BSA; Life Technologies), and 10 min with 1 µM Hoechst-33342-trihydrochloride. Three washes with PBS were performed after each staining step. Images were obtained using a Zeiss AxioObserver D1 microscope equipped with an LD A-Plan 20X/0.35 Ph1 objective.

*ROS production:* Neutrophils at 2 x $10^6$ cells/ml were incubated with 10 μM 2',7'-dichlorodihydrofluorescein diacetate ($H_2DCFDA$) in 1.5 ml siliconized tubes and incubated for 20 min at 37°C on an orbital shaker. Neutrophils were centrifuged at 500g for 8 min and cells were resuspended in original volume with $HBSS^{-/-}$. Neutrophils at 2 x $10^5$ in 100 μl were plated in a 96-well plate and exposed to a final concentration of 0 nM, 2.5 nM 25 nM and 250 nM of PMA (all conditions run in triplicate). Fluorescence was measured using an Infinite M200 (TECAN) plate reader with 495 nm excitation and 520 nm emission, every 15 min for 2 h, with incubation at 37°C and 5% $CO_2$ in-between.

*Phagocytosis:* Neutrophils were mixed in a 1:1 volume ratio with pHrodo™ Red *S. aureus* Bioparticles™ (Invitrogen) in a 96-well plate and centrifuged at 1200 rpm for 6 min. Fluorescence was measured using an Infinite M200 (TECAN) plate reader with 560 nm excitation and 585 nm emission every 15 min for 2 h, with incubation at 37°C and 5% $CO_2$ in-between. To exclude background signal, we also measured fluorescence of wells containing either HBSS, cells and pHrodo™ Red *S. aureus* Bioparticles.
